# Model-Based Evaluation of Colorectal Cancer Screening Effectiveness: Three Rounds of Multitarget Stool DNA Testing Versus One Colonoscopy

**DOI:** 10.64898/2026.01.30.26344467

**Authors:** Michael Dore, Derek W. Ebner, Vahab Vahdat, Chris Estes, A. Burak Ozbay, Victoria Foster, Paul J. Limburg

## Abstract

**Background:** Several colorectal cancer (CRC) screening modalities are guideline-recommended in the United States, yet they vary considerably in screening interval and real-world adherence. As a result, single-round test performance may not reflect cumulative effectiveness over time. This study compared the 10-year longitudinal outcomes of two CRC screening strategies— triennial next-generation multitarget stool DNA testing (ng mt-sDNA) and decennial screening colonoscopy.

**Methods:** This study used the validated, microsimulation-based Colorectal Cancer and Adenoma Incidence and Mortality (CRC-AIM) model to estimate 10-year cumulative outcomes for two guideline-recommended screening strategies: triennial ng mt-sDNA and decennial colonoscopy. Model inputs included test-specific performance and real-world adherence. Outcomes included CRC and precancerous lesions detected, CRC mortality reductions, and life-years gained (LYG). Sensitivity analyses examined the effects of varying both screening adherence and follow-up colonoscopy adherence.

**Results:** Over 10 years per 1,000 individuals offered screening, the ng mt-sDNA screening test detected 13% more precancerous lesions and 11% more CRC cases than colonoscopy, with a greater proportion of CRCs identified through screening rather than symptomatic detection. ng mt-sDNA resulted in greater CRC mortality reduction (33% vs 20%) and 62% more life-years gained, with consistent findings across sensitivity analyses.

**Conclusions:** With real-world adherence, screening with triennial ng mt-sDNA demonstrates superior cumulative effectiveness compared with decennial colonoscopy, driven by higher adherence and favorable longitudinal performance. These findings support the expanded use of noninvasive stool-based screening to reduce CRC mortality and alleviate capacity constraints associated with colonoscopy-based screening. Broader adoption of ng mt-sDNA may enhance population-level CRC prevention by increasing participation and improving early detection across the screening eligible population.

**Plain language summary:** Colorectal cancer screening tests are recommended at different time intervals and completed at different adherence rates in clinical practice. The analysis used a validated simulation model to compare 10-year outcomes of triennial next-generation multi-target stool DNA test (ng mt-sDNA) with a single colonoscopy, accounting for real-world screening and follow-up colonoscopy adherence. Our findings indicate that repeated ng mt-sDNA provides greater cumulative screening effectiveness than colonoscopy over a 10-year period.

## INTRODUCTION

Colorectal cancer (CRC) remains a major health challenge in the United States, ranking as the third most frequently diagnosed cancer and the second leading cause of cancer-related mortality. In 2025, it is projected to result in approximately 158,850 new diagnoses and 55,230 deaths.^1^ Although CRC is highly preventable through routine screening with identification and removal of precancerous lesions, it continues to have a substantial negative impact on public health. For adults ages 45 years or older and at average CRC risk, the U.S. Preventive Services Task Force (USPSTF) recommends several screening approaches, including the two most commonly utilized options: colonoscopy every 10 years and multitarget stool DNA (mt-sDNA) every 3 years.^2, 3^ Colonoscopy enables direct visualization of the colon and allows for the detection and removal of precancerous lesions during the same procedure. However, it is an invasive, resource-intensive, and capacity-constrained screening strategy that requires bowel preparation, sedation, and time away from work or other activities; factors that can negatively impact real-world adherence with guideline recommendations. As a result, the overall effectiveness of colonoscopy-based screening in routine practice is tempered by suboptimal patient participation. The next-generation mt-sDNA test (ng mt-sDNA; Cologuard Plus™ [Exact Sciences Corporation]) represents a noninvasive screening option that is completed at home without requiring any associated bowel preparation, dietary modification, or medication adjustments. Like the current mt-sDNA test, ng mt-sDNA uses the same patient support program, which has been shown to contribute to the strongest screening adherence.^4^ Screening performance for the ng mt-sDNA was demonstrated in the large, multicenter BLUE-C study (NCT04144738), with sensitivity for both CRC and advanced precancerous lesions (APLs) comparatively higher than an independent fecal immunochemical test (FIT).^5^ Previous studies reported that screening adherence rates are also higher for mt-sDNA compared to screening colonoscopy for initial screening and follow-up colonoscopy after positive stool test.^6-11^

Because overall CRC screening effectiveness is influenced by the combination of test characteristics, recommended interval, and patient participation over time, cumulative test performance across multiple screening rounds may differ substantially from single-test performance metrics. Accordingly, this modeling study uses a continuous-time agent-based microsimulation model to estimate and compare the cumulative detection of CRC and precancerous lesions over a 10-year time horizon using two strategies: triennial ng mt-sDNA screening and decennial colonoscopy, incorporating established test performance, screening intervals, and real-world adherence data. This analysis aims to inform clinical decision-making by clarifying how these strategies perform over time in routine screening populations.

## METHODS

### Model

For comparing the cumulative sensitivity and outcomes between ng mt-sDNA and colonoscopy, we used the Colorectal Cancer and Adenoma Incidence and Mortality (CRC-AIM) model, a microsimulation model incorporating CRC natural history as well as screening test characteristics to project long-term effects on CRC-related outcomes. CRC-AIM has undergone rigorous validation against US incidence and mortality data from the Surveillance, Epidemiology, and End Results (SEER) program^12^ and has been cross-validated against established Cancer Intervention and Surveillance Modeling Network (CISNET) models.^13^ The model has also been externally validated to other clinical trials such as United Kingdom Flexible Sigmoidoscopy (UKFSS).^14-16^ The natural history component, influenced by CISNET CRC-SPIN model,^17, 18^ simulates the adenoma–carcinoma sequence, including probabilistic adenoma initiation, growth, progression to preclinical cancer, transition to clinically detectable disease, and eventual survival or death. Screening interventions are modeled to reflect their ability to detect and remove adenomas or identify early-stage cancers, thereby reducing CRC mortality risk. A summary of the model is provided in Supplement 2.

### Screening Test Parameters

The model incorporated test-specific sensitivity and specificity, recommended screening intervals, adherence rates, and age-based eligibility criteria. Consistent with established CRC screening modeling frameworks, test performance was parameterized using adenoma sensitivity stratified by lesion size (<6 mm, 6–10 mm, and ≥10 mm), CRC sensitivity by disease stage, and overall test specificity (defined as the absence of colorectal neoplasia) (Table 1).

**Table 1.** Performance and adherence inputs for ng mt-sDNA and screening colonoscopy.

| Screening modality | Sensitivity |  |  |  | Specificity | Recommended screening interval | Adherence |  |
| --- | --- | --- | --- | --- | --- | --- | --- | --- |
|  | Precancerous lesions |  |  | CRC |  |  | Screening | Follow-up colonoscopy |
|  | <6 mm | 6–10 mm | ≥10 mm |  |  |  |  |  |
| ng mt-sDNA | 9.9% <sup>19</sup> | 17.8% <sup>19</sup> | 43.9% <sup>19</sup> | 95.3% <sup>19</sup> | 92.7% | 3 years <sup>2</sup> | 72% <sup>6</sup> | 77% <sup>7</sup> |
| colonoscopy | 75.0% <sup>20</sup> | 85.0% <sup>20</sup> | 95.0% <sup>20</sup> | 95.0% <sup>20</sup> | 86%* <sup>20</sup> | 10 years <sup>2</sup> | 38% <sup>8</sup> | — |

\*Specificity for colonoscopy reflects the detection of non-neoplastic lesions that may lead to unnecessary polypectomy which is associated with an increased risk of complications.

Performance estimates for ng mt-sDNA were derived from the FDA submission for the BLUE-C clinical trial (NCT04144738),^19^ a large retrospective study of more than 20,000 individuals in the United States, with Exact Sciences Corporation granting access and permissions to reuse the dataset for this research study. Because this is secondary data analysis using previously published and/or publicly available data, it does not require ethical approval as it does not involve the collection of new data from human participants. All data used were de-identified, and the original studies obtained all necessary informed consents and ethical approvals. Colonoscopy performance parameters were obtained from the published literature (Table 1).^20^ Based on these sensitivity and specificity inputs, screening outcomes were modeled as true positives (detection and removal of precancerous lesions or identification and treatment of CRC), true negatives (return to routine screening per the selected modality and interval), false positives (unnecessary diagnostic follow-up), and false negatives (missed adenomas or CRC).

The simulation estimated disease and screening outcomes over a 10-year horizon, initiating cohorts of 1 million average-risk individuals in the U.S. without diagnosed CRC at ages 45/55 and 65 years to represent commercially insured and Medicare-insured populations, respectively. Final results were weighted to reflect the U.S. population aged 45–54, 55-64, and 65–75 years using US Census data.^21^ Separated outcomes for commercial and Medicare populations are provided in Supplement 1.

### Strategy Assumptions

ng mt-sDNA was modeled at a three-year screening interval for 9-years (e.g. screening offered up to 3 times) while screening colonoscopy was modeled at a 10-year interval (one-time screening). Primary analyses assumed real-world adherence from published literature: screening colonoscopy (38%),^8^ ng mt-sDNA screening (72%),^6^ and follow-up colonoscopy after positive ng mt-sDNA (77%).^7^ Secondary analyses explored adherence scenarios in relative ±10 and ±20% increments for both screening and follow-up colonoscopy.

### Outcomes

Key outcomes included CRC cases detected, precancerous lesions detected, life-years gained (LYG) and reductions in CRC incidence and mortality compared with no screening per 1,000 individuals.

## RESULTS

Across a 10-year screening horizon, the ng mt-sDNA screening strategy resulted in 10.8 and 3.01 CRC cases and deaths compared to 9.7 and 3.61 for screening colonoscopy per 1,000 individuals offered screening (Figure 1). It was estimated that 7.4 (69%) of CRCs under the ng mt-sDNA strategy were detected by screening as compared to 1.8 (18%) for screening colonoscopy, a >275% increase. Detection of precancerous lesions was 13% higher with ng mt-sDNA as compared to screening colonoscopy (102 vs 90 detected per 1,000). Triennial ng mt-sDNA screening also resulted in a greater number of detected pre-symptomatic CRC cases; CRC mortality reduction as compared to no screening was 33% with ng mt-sDNA as compared to 20% for screening colonoscopy, a 68% relative increase. Estimated LYG were 5.1 per 1,000 with ng mt-sDNA as compared to 3.1 with screening colonoscopy (62% increase). Furthermore, LYG were 64% higher for commercially insured individuals (3.1 for ng mt-sDNA vs 1.9 for screening colonoscopy) and 61% higher for Medicare beneficiaries (9.4 vs 5.8 LYG).

**Figure 1.**
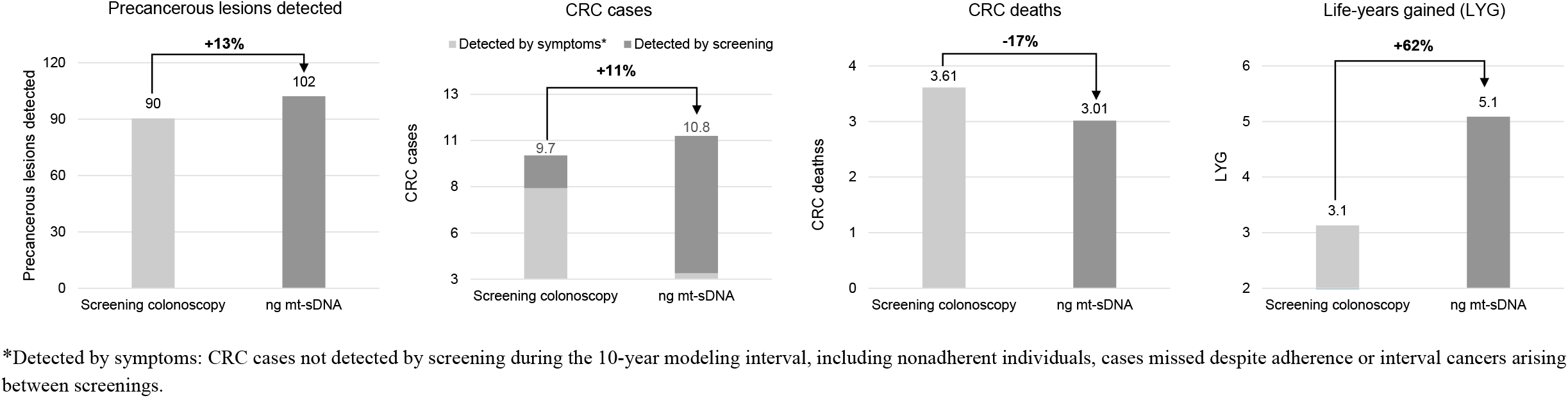
Comparative screening outcomes for ng mt-sDNA versus colonoscopy per 1,000 individuals screened over 10 years. *Detected by symptoms: CRC cases not detected by screening during the 10-year modeling interval, including nonadherent individuals, cases missed despite adherence or interval cancers arising between screenings.

When adherence to screening was concurrently varied for ng mt-sDNA and screening colonoscopy, LYG remained 56-70% higher with ng mt-sDNA (Table 2) and relative mortality reductions were 59-81% greater than screening colonoscopy. When follow-up adherence after a positive ng mt-sDNA was varied, LYG were 48-72% higher with ng mt-sDNA as compared to the base screening colonoscopy strategy, while relative mortality reductions were 55-76% higher (Table 2). In threshold analysis, screening colonoscopy required 43.4% adherence to match ng mt-sDNA in precancerous lesion detection, 63.1% for LYG, and 64.0% for CRC mortality reduction, and there was no adherence level at which colonoscopy could match ng mt-sDNA for pre-symptomatic CRC detection.

**Table 2.** Sensitivity analysis of screening outcomes under varying adherence levels for ng mt-sDNA and colonoscopy (per 1,000 individuals screened over 10 years)

| Modality | Screening adherence | Follow-up colonoscopy adherence | Precancerous lesions detected | Symptomatic CRC cases detected | Pre-symptomatic CRC cases detected | Total CRC cases detected | LYG | Mortality reduction |
| --- | --- | --- | --- | --- | --- | --- | --- | --- |
| ng mt-sDNA<br>Base screening adherence<br>Base follow-up adherence | 72% | 77% | 102 | 3.3 | 7.4 | 10.8 | 5.1 | 33% |
| ng mt-sDNA<br>+10% screening adherence<br>Base follow-up adherence | 79% | 77% | 109 | 2.8 | 7.8 | 10.6 | 5.4 | 35% |
| ng mt-sDNA<br>+20% screening adherence<br>Base follow-up adherence | 86% | 77% | 115 | 2.3 | 8.1 | 10.4 | 5.8 | 37% |
| ng mt-sDNA<br>-10% screening adherence<br>Base follow-up adherence | 65% | 77% | 95 | 3.9 | 7.0 | 10.9 | 4.7 | 31% |
| ng mt-sDNA<br>-20% screening adherence<br>Base follow-up adherence | 58% | 77% | 87 | 4.6 | 6.5 | 11.1 | 4.2 | 28% |
| ng mt-sDNA<br>Base screening adherence<br>+10% follow-up adherence | 72% | 85% | 102 | 3.1 | 7.5 | 10.6 | 5.3 | 34% |
| ng mt-sDNA<br>Base screening adherence<br>+20% follow-up adherence | 72% | 92% | 102 | 3.0 | 7.5 | 10.6 | 5.4 | 35% |
| ng mt-sDNA<br>Base screening adherence<br>-10% follow-up adherence | 72% | 69% | 102 | 3.5 | 7.3 | 10.8 | 4.8 | 32% |
| ng mt-sDNA<br>Base screening adherence<br>-20% follow-up adherence | 72% | 62% | 102 | 3.8 | 7.3 | 11.0 | 4.6 | 31% |
| Screening colonoscopy<br>Base screening adherence | 38% | — | 90 | 7.9 | 1.8 | 9.7 | 3.1 | 20% |
| Screening colonoscopy +10% screening adherence | 42% | — | 98 | 7.6 | 1.9 | 9.5 | 3.3 | 21% |
| Screening colonoscopy +20% screening adherence | 46% | — | 107 | 7.1 | 2.1 | 9.2 | 3.7 | 23% |
| Screening colonoscopy -10% screening adherence | 34% | — | 82 | 8.4 | 1.6 | 10.0 | 2.8 | 18% |
| Screening colonoscopy -20% screening adherence | 30% | — | 73 | 8.8 | 1.4 | 10.2 | 2.5 | 15% |

In a sensitivity analysis assuming perfect adherence for both strategies, outcomes were more comparable: mt-sDNA detected 9.8 CRC cases with a 43% mortality reduction, whereas screening colonoscopy detected 5.7 CRC cases with a 52% mortality reduction. Although perfect adherence is unlikely in practice, this scenario illustrates that differences between screening strategies narrow when adherence is not considered.

## DISCUSSION

Noninvasive stool-based tests have broadened access to CRC screening and have the potential to increase overall screening participation. Because the effectiveness of CRC screening at the population level depends not only on test performance but also on adherence and feasibility of implementation, identifying strategies that perform well under real-world conditions is critical. In this simulation, ng mt-sDNA screening conducted at 3-year intervals over a 10-year horizon was associated with greater CRC and advanced precancerous lesion detection, lower CRC mortality, and more life-years gained compared with screening colonoscopy. These findings highlight the potential clinical benefit of widely adopting noninvasive screening approaches, particularly in the context of constrained colonoscopy capacity. In the United States, more than 60 million adults are eligible for CRC screening,^22^ whereas annual colonoscopy capacity for screening purposes is estimated to be substantially lower.^23^ This imbalance has been further exacerbated by deferred procedures during the COVID-19 pandemic^24^ and by the expansion of the screening-eligible population following the 2021 USPSTF recommendation to lower the starting age to 45 years.^2^ Within this setting, scalable stool-based screening strategies may help prioritize colonoscopy resources for individuals most likely to benefit from diagnostic follow-up or therapeutic intervention, while reducing screening delays and enhancing population-level CRC prevention and mortality reduction.

Prior modeling studies have applied a cumulative sensitivity framework to enable evaluation of comparative effectiveness between screening modalities with differing test intervals.^25^ The cumulative sensitivity and specificity can be calculated as *(1 – (1 – sensitivity)*^*x*^ *) and specificity*^*x*^, where *x* represents the number of screening rounds. This method serves as a proxy for comparing the cumulative performance of the tests independent of observed adherence differences. We took further steps with calculating the age weighted cumulative performance based on US census data, as proposed by Shaukat et al.^26^ Applying this framework, the cumulative sensitivity of ng mt-sDNA over nine years of recommended use is estimated to be 100% and 81% for age-weighted CRC and APL sensitivity, and 100% and 82% for unweighted sensitivity, respectively (Supplement 1). While informative, this analysis is limited as test results are conditionally independent given the true disease state—such that lesion detection is equally likely at each screening round.

Additionally, long-term real-world comparative studies of clinical outcomes after multiple rounds of CRC screening remain limited. However, one retrospective cohort study evaluated CRC incidence and mortality among individuals screened with stool-based tests compared with those undergoing colonoscopy using data from the TriNetX database.^27^ Adults aged 45–80 years were categorized by screening modality (gFOBT or mt-sDNA) and matched 1:1 to a colonoscopy cohort using propensity score methods, resulting in a matched population of more than 371,000 patients. In this analysis, mt-sDNA screening was associated with higher CRC detection rates and a 51% reduction in the odds of CRC-related mortality compared with colonoscopy. These findings are consistent with the outcomes observed in the present simulation, supporting the potential population-level benefits of stool-based screening when implemented in real-world settings.

This analysis has several limitations. First, other CRC screening options used in the United States warrant further evaluation. FIT is another guideline-recommended stool-based test; however, because prior studies have demonstrated superior test performance and effectiveness of mt-sDNA compared with FIT,^5, 28^ further evaluation of FIT was not emphasized. Second, the modeling analyses were intentionally limited to a 10-year screening period rather than lifetime screening. Although longer-term models may more fully capture the cumulative benefits, burdens, and potential harms of screening, such lifetime analyses have been previously reported.^20, 29^ Moreover, from the perspective of certain payers and health systems—where patient enrollment and follow-up may occur over shorter time horizons—a 10-year analytic framework provides complementary and decision-relevant information regarding intermediate clinical outcomes and resource utilization. As reported, the more favorable stage distribution at diagnosis and the higher proportion of pre-symptomatic CRCs detected with ng mt-sDNA may suggest the potential for additional downstream benefits over a longer screening period, including improved survival and lower treatment costs, as cancers diagnosed at earlier stages are generally more amenable to curative therapy. Also, this analysis did not include a formal cost-effectiveness evaluation, as such analyses require longer time horizons to more accurately estimate costs and utilities, as well as longitudinal adherence modeling to capture individualized screening behaviors over time. Third, adherence to CRC screening may vary across populations according to race and ethnicity, socioeconomic status, and geographic access (rural vs urban). Sensitivity analyses addressing a range of adherence assumptions demonstrated that ng mt-sDNA consistently maintained advantages over colonoscopy across key clinical outcomes. However, the model did not incorporate dynamic, time varying adherence or adherence conditional on prior screening behavior. Because follow-up after a positive ng mt-sDNA test may have an even higher diagnostic yield (i.e., detection rate for CRCs and precancerous lesions), this assumption likely results in a conservative estimate of the comparative benefits of a ng mt-sDNA versus screening colonoscopy strategy. Additionally, the current model does not include the sessile serrated pathway (SSP). Because SSPs contribute meaningfully to CRC development and vary in detectability across screening modalities, future research should incorporate SSP-specific incidence, progression, and detection parameters to more fully evaluate potential differences in clinical effectiveness.

In summary, the findings from this microsimulation modeling study suggest that a shift toward broader adoption of noninvasive stool-based screening strategies may be an effective option, considering the colonoscopy capacity constraints and low adherence to such test. When longitudinal screening performance and real-world adherence to screening is considered, repeat ng mt-sDNA testing suggests cumulative effectiveness for colorectal cancer screening that is more favorable to decennial colonoscopy. Future research should continue to evaluate longer-term outcomes, cost-effectiveness, and strategies to optimize follow-up after positive tests and to maximize the benefits of CRC screening programs.

## Data Availability Statement

All data produced in the present study are available upon reasonable request to the authors.

