## Supplement 1 for "Model-Based Evaluation of Colorectal Cancer Screening Effectiveness: Three Rounds of Multitarget Stool DNA Testing Versus One Colonoscopy"

### Supplement 1. Supporting Analyses

#### Supplemental Table 1. Cumulative performance* of ng mt-sDNA over three consecutive rounds†

| | Repeat screening with ng mt-sDNA | CRC sensitivity | APL sensitivity§ | Specificity¶ |
|---|---|---|---|---|
| **Age weighted cumulative performance‡** | 1 round | 97% | 42% | 94% |
| | 2 consecutive rounds | 100% | 66% | 88% |
| | 3 consecutive rounds | 100% | 81% | 83% |
| **Unadjusted cumulative performance** | 1 round | 95% | 43% | 93% |
| | 2 consecutive rounds | 100% | 68% | 86% |
| | 3 consecutive rounds | 100% | 82% | 80% |

Abbreviations: APL, advanced precancerous lesion; CRC, colorectal cancer; ng mt-sDNA, next-generation multi-target stool DNA.

*Performance estimates for ng mt-sDNA derived from the primary effectiveness cohort for participants aged 45-84 years from the FDA submission for the BLUE-C clinical trial (NCT04144738).[1]

†Estimates assume that test results are independent from one testing round to the next and that no disease progression occurs.

‡FDA submission performance from the BLUE-C clinical trial (NCT04144738)[1] for participants aged 45-84 years weighted to 2020 US Census population estimates by age group.[2]

§APL performance includes 6 lesions <5 mm, 71 lesions 5-9 mm, and 1,884 lesions ≥10 mm.[1]

¶Specificity based on category 6 (negative: no adenocarcinoma of the colorectum, no adenomas or SSA/SSP).[1]

1. US Food and Drug Administration. Including data from the primary effectiveness cohort reviewed and approved by the FDA, as summarized in PMA P230043: FDA Summary of Safety and Effectiveness Data. Multi-Target Stool DNA (mt-sDNA) Based Colorectal Cancer Screening Test, https://www.accessdata.fda.gov/cdrh_docs/pdf23/P230043B.pdf (2024).
2. Blakeslee L, Caplan Z, Meyer JA, et al. Age and Sex Composition: 2020 Census Briefs (2020 Census Briefs C2020BR-06). https://www2.census.gov/library/publications/decennial/2020/census-briefs/c2020br-06.pdf. 2023.

**Supplemental Table 2. Screening outcomes for ng mt-sDNA and colonoscopy among commercial and Medicare populations (per 1,000 individuals screened over 10 years)**

| Modality | Screening adherence | Follow-up colonoscopy adherence | Precancerous lesions detected | Symptomatic CRC cases detected | Pre-symptomatic CRC cases detected | Total CRC cases detected | LYG | Mortality reduction |
|---|---|---|---|---|---|---|---|---|
| **ng mt-sDNA (commercial population)** | 72% | 77% | 87 | 2.2 | 5.0 | 7.2 | 3.1 | 35% |
| **Screening colonoscopy (commercial population)** | 38% | – | 79 | 5.4 | 1.1 | 6.5 | 1.9 | 21% |
| **ng mt-sDNA (Medicare population)** | 72% | 77% | 134 | 5.7 | 12.7 | 18.4 | 9.4 | 32% |
| **Screening colonoscopy (Medicare population)** | 38% | – | 114 | 13.3 | 3.2 | 16.5 | 5.8 | 19% |

Abbreviations: CRC, colorectal cancer; LYG, life-years gained; ng mt-sDNA, next-generation multi-target stool DNA.
