## Supplement 2 for "Model-Based Evaluation of Colorectal Cancer Screening Effectiveness: Three Rounds of Multitarget Stool DNA Testing Versus One Colonoscopy"

**Supplement 2: Colorectal Cancer Adenoma Incidence and Mortality (CRC-AIM) Model**

### A. CRC-AIM model structure

CRC-AIM models the natural history of colorectal cancer (CRC) using an adenoma-carcinoma sequence. The natural history of CRC is comprised of five components: (1) adenoma generation; (2) adenoma growth; (3) transition from adenoma to preclinical cancer; (4) transition from preclinical cancer to clinically detectable cancer (i.e., sojourn time); and (5) survival. Individuals may develop one or more adenomas, which may transition into a preclinical cancer as it grows. A preclinical cancer may ultimately transition into a clinically detectable cancer, and then lead to CRC-specific mortality. CRC-AIM does not model CRCs that occur through sessile serrated pathway (SSP), which is a major limitation of our model. Approximately 14% - 30%[3-5] of CRCs are estimated to arise from sessile serrated lesions and polyps, which develop mainly via the CpG island methylation pathway.[6, 7] In addition, the CRC screening modalities vary substantially in the detection of SSPs, since serrated polyps are less likely to bleed compared to adenoma.[4] Also, our model does not represent the CRC-related events experienced by individuals who are at high risk such as patients with inflammatory bowel disease (e.g., Crohn's disease, ulcerative colitis) and those with a personal or family history of CRC. This is because such patients are likely to have a different natural history of CRC and require different screening/surveillance strategies than average-risk individuals.

The ColoRectal Cancer Simulated Population Incidence and Natural history (CRC-SPIN) model (one of three CISNET CRC models) inspired the development of CRC-AIM, and the models share features (as obtained or derived from publicly available sources).[8, 9]

However, in addition to some structural differences, almost all the parameter values differ between CRC-AIM and CRC-SPIN as explained in detail in Supplemental Section B.

In the remainder of this section, we provide details of each of the five subcomponents of CRC-AIM.

*1. Adenoma generation*

The risk of developing adenoma is assumed to depend on individuals' sex, age and baseline risk, with individuals aged ≤20 years not at risk for adenoma development.[10] The risk of generation an adenoma is governed by a non-homogenous Poisson process based on baseline risk (varying by individuals), sex, and age. We introduce the following notation:

- $r_i(a)$: risk of developing an adenoma for individual $i$ at age $a$;
- $I_F(i)$:indicator function, which is equal to 1 when individual $i$ is female and 0 otherwise;
- $I_{[a_0,\infty)}(a)$: indicator function that is equal 1 if the invidual's age $a \in [a_0, \infty)$;
- $\beta_j$:regression coefficients of the log-linear model.

The functional represention of the adenoma risk model may then be expressed as:

$$\ln(r_i(a)) = \beta_{0i} + \beta_1 I_F(i) + \beta_2 I_{[20,\infty)}(a)\min\{a-20,30\} + \beta_3 I_{[50,\infty)}(a)\min\{a-50,10\} + \beta_4 I_{[60,\infty)}(a)\min\{a-60,10\} + \beta_5 I_{[70,\infty)}(a)(a-70)$$

Upon the generation of an adenoma, its location is determined via a multinomial distribution informed by autopsy studies[11-19] as derived in Rutter et al.[9] (Supplemental Table 1).

**Supplemental Table 1. Multinomial distribution used for assigning the location of new adenomas**

| Site | Rectum | Distal colon | | Proximal colon | | |
|---|---|---|---|---|---|---|
| | | Sigmoid | Descending | Transverse | Ascending | Cecum |
| **Probability** | 0.09 | 0.24 | 0.12 | 0.24 | 0.23 | 0.08 |

2. *Adenoma growth*

The diameter in millimeters (mm) of an adenoma $j$ at time $t$, measured after its initiation, in individual $i$ is modeled using the Richard's growth model:[20]

$$d_{ij}(t) = d_{max}\left[1 + \left(\left(\frac{d_{min}}{d_{max}}\right)^{\frac{1}{p}} - 1\right)e^{-\lambda_{ij}t}\right]^p$$

where $p$ represents the unknown parameter of the growth model, $\lambda_{ij}$ represents the growth rate of adenoma $j$ in individual $i$, $d_{min}$ and $d_{max}$ represent the minimum and maximum diameter. The diameter of an adenoma is confined between 1 mm and 50 mm, so that $d_{min} = 1\ mm$ and $d_{max} = 50\ mm$.

The growth rate $\lambda_{ij}$ is determined stochastically by first sampling the time to reach 10 mm in diameter, denoted as $t_{10mm}$, which is assumed to follow the Fréchet distribution with scale parameter $s_l$ and shape parameter $\alpha_l$ ($l \in \{\text{colon}, \text{rectum}\}$). The cumulative distribution function (CDF) at time $t$ after the initiation of an adenoma located at $l$ is given by

$$F_l(t) = \exp\left[-\left(\frac{t}{s_l}\right)^{-\alpha_l}\right]$$

Once $t_{10mm}$ is sampled from the CDF, the growth rate can be determined by solving the growth model using $d_{ij}(t_{10mm}) = 10$, leading to:

$$\lambda_{ij} = -\frac{1}{t_{10mm}}\ln\frac{\left(\frac{10}{d_{max}}\right)^{\frac{1}{p}} - 1}{\left(\frac{d_{min}}{d_{max}}\right)^{\frac{1}{p}} - 1}$$

3. *Transition from adenoma to preclinical cancer*

The cumulative transition probability of progressing from adenoma to preclinical cancer was modeled using a lognormal cumulative distribution function based on sex, size and age at adenoma initiation.[21, 22] For an adenoma that is initiated at age $a$ located in $l \in \{\text{colon}, \text{rectum}\}$ of an individual whose sex is $s \in$ {male, female}, the probability of transition to preclinical cancer at or before diameter $d$ is given by:

$$P_{ls}(d, a) = \Phi\left(\frac{\ln(\gamma_{1ls} d) + \gamma_{2ls}(a - 50)}{\gamma_3}\right)$$

where $\Phi(\cdot)$ denotes the standard normal CDF. Following Rutter et al.,[9] $\gamma_3$ is set to be 0.5. The annual transition probability from year $t$ to $t + 1$ may be expressed as

$$\frac{P_{ls}(d_{t+1}, a) - P_{ls}(d_t, a)}{1 - P_{ls}(d_t)}$$

where $d_t$ represents the diameter of the adenoma at year $t$.

4. *Transition from preclinical cancer to clinically detectable cancer (sojourn time)*

For colon adenomas, the sojourn time is modeled using a Weibull distribution with shape parameter $k$ and location-specific scale parameter $\lambda_c$ where the survival function is given by:

$$S_c(t) = \exp\left(-\left(\frac{t}{\lambda_c}\right)^k\right)$$

Assuming proportional hazards between rectal and colon adenomas with the following hazard ratio (HR),

$$\mathrm{HR} = \exp(\alpha)$$

Rectal adenomas also follow Weibull distribution with the following survival distribution

$$S_r(t) = \exp\left(-\left(\frac{t}{\lambda_c\, exp(\alpha)^{-\frac{1}{k}}}\right)^k\right)$$

Here, $\lambda_r = \lambda_c \exp(\alpha)^{-\frac{1}{k}}$. In summary, the sojourn time model for both colon and rectal adenomas has a total of three parameters: $\lambda_c$, $k$, and $\alpha$.

a. *CRC stage at clinical detection*

When a preclinical cancer becomes clinically detectable (i.e., at the end of sojourn time), CRC-AIM first attributes the AJCC stage (5$^{th}$ edition) at clinical detection using a multinomial distribution, stratified by anatomic subsite location (proximal colon, distal colon, and rectum), age-group at diagnosis, and sex. Proximal colon anatomic subsite includes cecum, ascending, hepatic flexure, transverse colon, and splenic flexure. Distal colon consists of descending colon and sigmoid colon. Finally, rectosigmoid colon and rectum are included in rectum. The distributions were derived from SEER 1975-1979 data. Since AJCC staging was not recorded prior to 1988, we adopted the methodology introduced by Schrag[23] for estimating the cancer stages by combining site-specific surgery codes and the staging codes (Supplemental Figure 1).

**Supplemental Figure 1. Distribution of stage at clinical detection by age, sex, and location according to SEER 1975-1979 data**

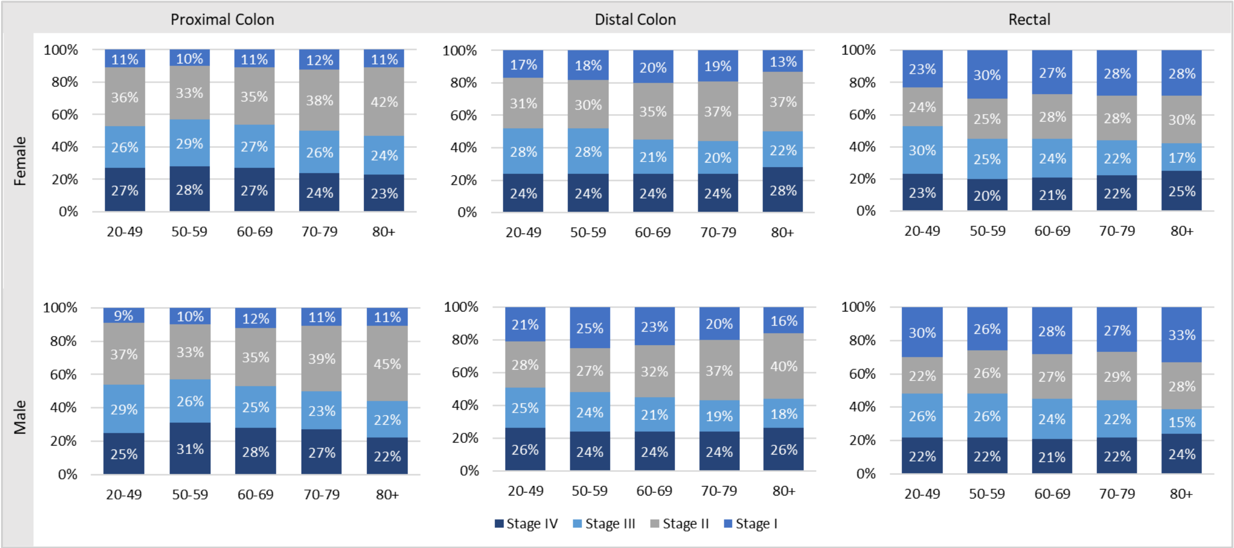


*b. CRC size at clinical detection*

The size at clinical detection, conditional on location and stage at clinical detection, is assigned using SEER 2010-2015 data, limited to cases diagnosed at ages 20-50. There has been a difference between SEER 2010-2015 data and SEER 1975-1979 data for the same stage (AJCC 5$^{th}$ edition), age group, and anatomic location. Depending on location, stage I tumors in the 2010-2015 data were 10-15 mm smaller, on average, than the stage I tumors in the 1975-1979 data. There was no effect of age on tumor size at diagnosis in the 1975-1979 data, nor was there a difference in stage I tumor size between patients younger than 50 years compared with patients 50 years or older. This suggests that in modern times, even without screening, stage I tumors may be diagnosed when they are smaller.

The true tumor size at diagnosis by stage and location was assumed to follow a gamma distribution. Within the 8,577 cases retrieved from SEER 2010-2015, tumors larger than 150 mm (n = 29, 0.5%) were set to equal to 150 mm and tumor sizes coded as more than 900 mm (n = 1,820) were excluded. Tumors in the appendix or unspecified locations of the large intestine were excluded (n = 992). After exclusions, 5,910 cases remained for modeling. Right censoring (i.e., sizes greater than 98 mm that are recorded as 98 mm as in the SEER 1975-1979 data) is more likely to skew distribution estimates than interval censoring (i.e., rounding of tumor size data). Because SEER 2010-2015 data were not right-censored, gamma distributions were fit to the data while ignoring the interval censoring using maximum likelihood (Supplemental Table 2).

**Supplemental Table 2. Estimated parameters of gamma distribution by stage and location that is used to assign tumor size in CRC-AIM**

| Location | Stage I | | Stage II | | Stage III | | Stage IV | |
|---|---|---|---|---|---|---|---|---|
| | Shape | Rate | Shape | Rate | Shape | Rate | Shape | Rate |
| **Distal Colon** | 1.5545 | 0.0639 | 5.5037 | 0.0998 | 4.3892 | 0.0919 | 5.3333 | 0.1050 |

| | | | | | | | | |
|---|---|---|---|---|---|---|---|---|
| **Proximal Colon** | 1.6695 | 0.0552 | 5.1046 | 0.0833 | 4.4307 | 0.0796 | 4.9185 | 0.0836 |
| **Rectum** | 1.1040 | 0.0645 | 3.7173 | 0.0777 | 3.8285 | 0.0763 | 3.6448 | 0.0641 |

5. *CRC survival*

CRC survival is based on parametric models with sex and age at diagnosis as covariates for each stage and location (colon vs rectum) fitted to SEER-reported cause-specific survival. A 7% hazard reduction was applied based on 5-year cause-specific relative survival between periods 2000-2003 and 2010-2019 from SEER, for cases diagnosed after 2000 to replicate the recent improvement in CRC-specific survival.[24] Other-cause mortality by age and birth-year cohorts were based on the U.S. life tables.[25]

Once an individual is diagnosed with CRC, they are assigned to CRC-specific survival, which is estimates as follows. First, we fit five parametric regression models (Weibull, lognormal, exponential, Fréchet, and loglogistic) to the cause-specific survival data from SEER 2000-2003 for each cancer stage and location (colon vs rectum). Each regression model included two sets of covariates: age at diagnosis (20-49, 50-59, 60-69, 70-79, and ≥80 years) and sex (1 if female and -1 otherwise). We applied right-censoring as appropriate. Any subject with a time of death that was reliably recorded as 0 months was recoded to 0.5 months to prevent model-fitting issues. Among the five regression models, we chose the one based on the smallest corrected Akaike information criterion (AICc) value. Statistical significance of age and sex was based on the Wald test, and the final model (presented in Supplemental Table 3) was chosen by refitting the parametric regression models while excluding any covariate that was not statistically significant. Similar analyses were performed for the cause-specific CRC survival distributions based on the SEER 1975-1979 data, which were used for comparing natural history outcomes against those from the CISNET models (data not shown).

**Supplemental Table 3. Selected parametric regression model by stage and location based on SEER 2000-2003 cause-specific survival**

| Location | Parameters | Stage I | Stage II | Stage III | Stage IV |
|---|---|---|---|---|---|
| **Colon** | **Selected distribution** | **Weibull** | **Log-normal** | **Log-normal** | **Log-normal** |
| | Intercept | 5.8797 | 4.5013 | 2.5361 | -0.3766 |
| | Age effect, age ∈ [20,50) | 1.6097 | 0.6193 | 0.5416 | 0.6531 |
| | Age effect, age ∈ [50,60) | 0.6499 | 0.3778 | 0.4263 | 0.3190 |
| | Age effect, age ∈ [60,70) | -0.0115 | 0.3243 | 0.2067 | 0.0861 |
| | Age effect, age ∈ [70,80) | -0.4863 | -0.2226 | -0.1463 | -0.2930 |
| | Age effect, age ≥ 80 | -1.7619 | -1.0987 | -1.0283 | -0.7653 |
| | Main sex effect | 0.1321 | 0.1825 | 0 | 0 |
| | Shape parameter | 0.6991 | 2.8316 | 2.2040 | 1.5583 |

| Rectum | Selected distribution | Log-logistic | Log-normal | Log-normal | Log-normal |
|---|---|---|---|---|---|
| | Intercept | 4.2475 | 3.4680 | 2.3558 | -0.1707 |
| | Age effect, age ∈ [20,50) | 0.4703 | 0.9608 | 0.4700 | 0.5883 |
| | Age effect, age ∈ [50,60) | 0.7033 | 0.4976 | 0.3899 | 0.5408 |
| | Age effect, age ∈ [60,70) | 0.1593 | 0.1146 | 0.3039 | 0.0489 |
| | Age effect, age ∈ [70,80) | -0.2561 | -0.3914 | -0.1072 | -0.3437 |
| | Age effect, age ≥ 80 | -1.0768 | -1.1817 | -1.0567 | -0.8343 |
| | Main sex effect | 0 | 0 | 0 | 0 |
| | Shape parameter | 0.9026 | 2.1874 | 1.7519 | 1.4349 |

Abbreviations: SEER, Surveillance, Epidemiology, and End Results.

#### 6. *List of calibrated natural history parameters*

Twenty-three directly unobservable parameters governing the natural history of CRC (Supplemental Table 4), were estimated using calibration for CRC-AIM. Parameter calibration began with a plausible range for each parameter, informed by CRC-SPIN.[8, 9] The initial plausible range was then supplemented using our calibration process.

**Supplemental Table 4. Unknown parameters of CRC-AIM natural history model (adapted from Vahdat et al.[26])**

| Unknown Parameter | Plausible Range | Best parameter value selected by calibration |
|---|---|---|
| **Adenoma generation** | | |
| Baseline log-risk, $\beta_0$ | $\beta_0 \sim TN_{[-7,-5]}(-6.3,\ 0.4)$ | $-5.661$ |
| Standard deviation of baseline log-risk, $\sigma_0$ | $\sigma_0 \sim TN_{[1,2]}\ (1.1,\ 0.2)$ | 1.270 |
| Sex effect, $\beta_1$ | $\beta_1 \sim TN_{[-0.5,-0.1]}(-0.5, 0.1)$ | $-0.384$ |
| Age effect (ages 20- <50), $\beta_2$ | $\beta_2 \sim TN_{[0.03,0.07]}(0.045, 0.007)$ | 0.039 |
| Age effect (ages 50- <60), $\beta_3$ | $\beta_3 \sim TN_{[0.01,0.05]}(0.03, 0.01)$ | 0.023 |
| Age effect (ages 60- <70), $\beta_4$ | $\beta_4 \sim TN_{[-0.01,0.05]}(0.03, 0.01)$ | 0.020 |
| Age effect (ages ≥ 70), $\beta_5$ | $\beta_5 \sim TN_{[-0.02,0.03]}(0.03, 0.03)$ | $-0.018$ |
| **Adenoma growth (time to 10 mm)** | | |
| Scale (colon), $s_c$ | $s_c \sim U(10.7,40)$ | 24.364 |
| Shape (colon), $\alpha_c$ | $\alpha_c \sim U(0.5,4)$ | 1.388 |
| Scale (rectum), $s_r$ | $s_r \sim U(5,20)$ | 6.734 |
| Shape (rectrum), $\alpha_r$ | $\alpha_r \sim U(2,5)$ | 3.601 |
| **Adenoma growth (Richard's growth model)** | | |
| Shape parameter, $p$ | $p \sim TN_{[0.5,3.2]}(1.0,\ 0.5)$ | 0.710 |
| **Transition from adenoma to cancer** | | |
| Size (male, colon), $\gamma_{1cm}$ | $\gamma_{1cm} \sim U(0.02,0.06)$ | 0.040 |
| Age at initiation (male, colon), $\gamma_{2cm}$ | $\gamma_{2cm} \sim U(0.0,0.02)$ | 0.016 |

| | | |
|---|---|---|
| Size (male, rectum), $\gamma_{1rm}$ | $\gamma_{1rm} \sim U(0.02,0.07)$ | 0.039 |
| Age at initiation (male, rectum), $\gamma_{2rm}$ | $\gamma_{2rm} \sim U(0.0,0.02)$ | 0.004 |
| Size (female, colon), $\gamma_{1cf}$ | $\gamma_{1cf} \sim U(0.02,0.05)$ | 0.043 |
| Age at initiation (female, colon), $\gamma_{2cf}$ | $\gamma_{2cf} \sim U(0.0,0.02)$ | 0.014 |
| Size (female, rectum), $\gamma_{1rf}$ | $\gamma_{1rf} \sim U(0.02,0.055)$ | 0.035 |
| Age at initiation (female, rectum), $\gamma_{2rf}$ | $\gamma_{2rf} \sim U(0.0,0.02)$ | 0.010 |
| **Sojourn time** | | |
| Scale (colon), $\lambda_c$ | $\lambda_c \sim U(3.0,5.0)$ | 4.683 |
| Shape (colon and rectum), $k$ | $k \sim U(2.0,5.0)$ | 3.620 |
| Log-hazard ratio, $\alpha$ | $\alpha \sim U(-1.0,1.0)$ | $-0.018$ |

$TN_{[a,b]}(\mu,\ \sigma)$ represents a truncated normal distribution with mean μ and standard deviation σ over the domain [a,b]. U(a,b) represents a uniform distribution with domain (a,b).

### B. Differences between CRC-AIM and CRC-SPIN

While CRC-AIM was inspired by CRC-SPIN, there are several differences. Almost all the estimated parameters between CRC-AIM and CRC-SPIN differ as the two models use different approaches for model calibration. Namely, while CRC-SPIN used a Bayesian calibration method, we utilize a machine learning-based empirical calibration approach. The differences in estimated parameters led to different predicted outcomes by the models (e.g., sojourn time, dwell time, adenoma size distribution, life-years gained, incidence reduction, and mortality reduction), further highlighting the importance of including structural or parametric sensitivity analysis within natural history modeling to ensure robust conclusions.

In addition to the differences in parameter values, there are three major structural differences between the two models. First, CRC-AIM uses yearly cycles for adenoma generation and transition into preclinical cancer, as opposed to a continuous-time model employed by CRC-SPIN. While the use of yearly cycles precludes the generation of multiple adenomas or transition from adenoma to clinical CRC within the same year, such events are rare as quantified by the transition probabilities reported before.[27] Second, CRC-AIM models survival from CRC using cause-specific survival, rather than the relative survival estimates from an analysis of SEER 1975-2003 data.[9] A presentation by NCI notes the limitations associated with using relative survival and suggests the use of cause-specific survival for accurate representation of post-diagnosis survival.[28] Additionally, CRC-AIM considers the improvement in survival since 2003 through advancements in cancer treatments by applying a 7% hazard reduction, which is estimated using the 5-year survival between periods 2000-2003 and 2010-2019 from SEER. No survival improvement was modeled in CRC-SPIN. Third, the CRC size and stage estimation differs between the two models. The stage at clinical detection in CRC-AIM is modeled as a function of anatomic subsite location (proximal colon, distal colon, and rectum), age-group at diagnosis, and sex. CRC-SPIN only included age and sex in modeling this variable. CRC-AIM models the size at clinical detection based on location and stage at clinical detection using SEER 2010-2015 data, limited to cases diagnosed at ages 20-50, where the rationale has been provided in the main manuscript. On the other hand, CRC-SPIN used 1975-1979 SEER data for this purpose. CRC-AIM models stage-shift due to screening by randomly assigning a stage at screening detection based on the distribution of stage at clinical detection but imposing the restriction that the newly sampled stage must not be worse than the stage at clinical detection. The modeling of stage shift in CRC-SPIN is not reported in detail.

### C. Summary of model inputs and assumptions

Supplemental Table 5 summarizes the methods, inputs, and assumptions employed by the CRC-AIM.

**Supplemental Table 5. Summary of CRC-AIM model components**

| Category | Component | Method | Source |
|---|---|---|---|
| **Model structure** | Model structure | Microsimulation model | |
| **Population characteristics** | Other-cause mortality | 2017 U.S. life table | Arias and Xu[29] |
| **Natural history** | Adenoma generation, growth, and transition to preclinical cancer | Calibrated to published literature data. | Corley et al.,[30] Pickhardt et al.,[31] Imperiale et al.,[32] Lieberman et al.,[33] Church[34] |
| | Adenoma location | Six locations (rectum, sigmoid, descending, transverse, ascending, cecum). | 9 autopsy studies[11-19] as derived in Rutter et al.[9] |
| | CRC incidence | Calibrated to SEER 1975-1979 incidence. | SEER[35] |
| | Symptomatic detection (stage at diagnosis) | A multinomial distribution derived from SEER 1975-1979 data. | SEER[35] |
| | Symptomatic detection (size at diagnosis) | A gamma distribution conditional on the stage at diagnosis, derived from SEER 2010-2015 data. | SEER[35] |
| | CRC survival | Parametric linear regression model derived from cause-specific survival from SEER 2000-2003. A 7% reduction in hazard, estimated using the 5-year cause-specific relative survival between periods 2000-2003 and 2010-2019 from SEER, is applied to cases diagnosed post 2000. | SEER[35] |
| **Screening analyses** | CRC incidence | An incidence rate ratio of 1.19 is applied to the baseline risk of generating adenomas. | Siegel et al.,[36] Knudsen et al.[37] |
| | Cohort of interest | Average-risk 40-year-old individuals in the U.S. (1980 birth cohort) who are previously unscreened for colorectal cancer and free of diagnosed colorectal cancer. | Knudsen et al.[37] |
| | Stage shift | Stage at screening detection is resampled from the stage at clinical diagnosis distribution and | |

| | | | |
|---|---|---|---|
| | | limited to no worse than the stage at symptomatic diagnosis. | |
| | Screening performance (sensitivity and specificity) | Varies by screening modality. Sensitivity is based on the cancer or the size of adenoma (1 to < 6 mm, 6 to < 10 mm, and ≥ 10 mm). | Knudsen et al.[37] |
| | Reach | The reach of colonoscopy linearly increases from rectum (100%) to cecum (95%). | Knudsen et al.[37] |
| | Screening complications | No complications assumed for stool tests. For colonoscopy with polypectomy, age-specific risks of serious gastrointestinal events, other gastrointestinal events, and cardiovascular events are included. | Knudsen et al.[37] |
| | False-positive test management | Resume screening with original modality 10 years after the false-positive test. | Knudsen et al.[37] |

Abbreviations: CRC, colorectal cancer; CRC-AIM, Colorectal Cancer-Adenoma Incidence and Mortality model; FIT, fecal immunochemical test; mt-sDNA, multi-target stool DNA; SEER, Surveillance, Epidemiology, and End Results.

### D. CRC-AIM natural history validation

We compared model outcomes to SEER 1975-1979 CRC incidence per 100,000 to validate the natural history of CRC. SEER includes the most comprehensive population-based nationwide CRC data before widespread CRC screening in the US, thus forming critical input for natural history model development. These data have been utilized by several other models of CRC in the US (including CISNET CRC models).[38-41] The primary targets were supplemented by data from studies by Corley et al.[30] and Pickhardt et al.,[31] which reported adenoma prevalence and distribution by size based on a large sample of asymptomatic patients.[30, 31]

Age-specific CRC incidence as reported by SEER's 1975-1979 data was matched by CRC-AIM (Supplemental Figure 2) along with adenoma prevalence reported by the autopsy studies[37] (Supplemental Figure 3). The dwell time and sojourn time estimated by CRC-AIM were 20.3 years and 4.1 years, respectively, both of which fall within the estimated values from the literature[42-44] and CISNET models (Supplemental Figure 4). Additional calibration targets are compared in Supplemental Table 6.

**Supplemental Figure 2. CRC-AIM and CISNET predictions of colorectal cancer cases per 100,000 people by age (adapted from Knudsen et al.[37])**

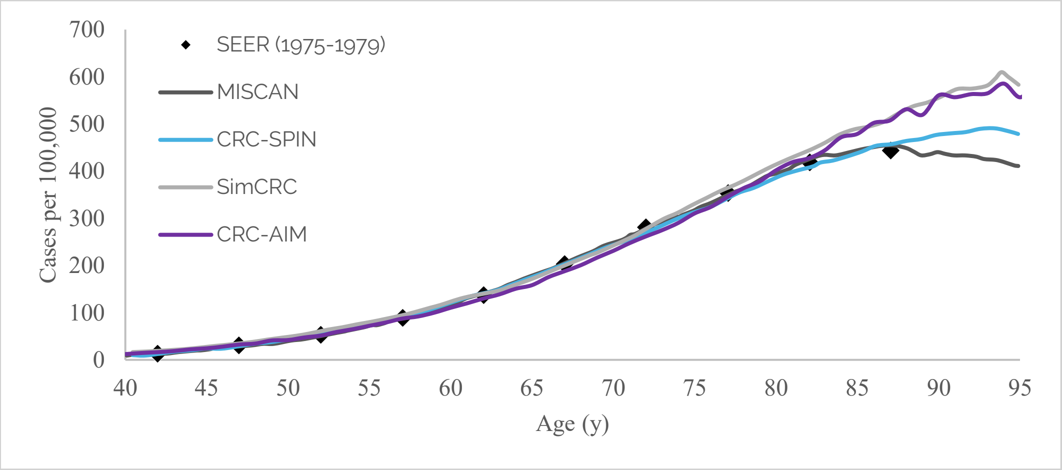


Abbreviations: CRC-AIM, Colorectal Cancer-Adenoma Incidence and Mortality model; CRC-SPIN, ColoRectal Cancer Simulated Population Incidence and Natural history model; MISCAN, MIcrosimulation SCreening Analysis; SEER, Surveillance, Epidemiology, and End Results; SimCRC, Simulation Model of Colorectal Cancer.

**Supplemental Figure 3. CRC-AIM predicted adenoma prevalence by age against autopsy data (adapted from Knudsen et al.[37])**

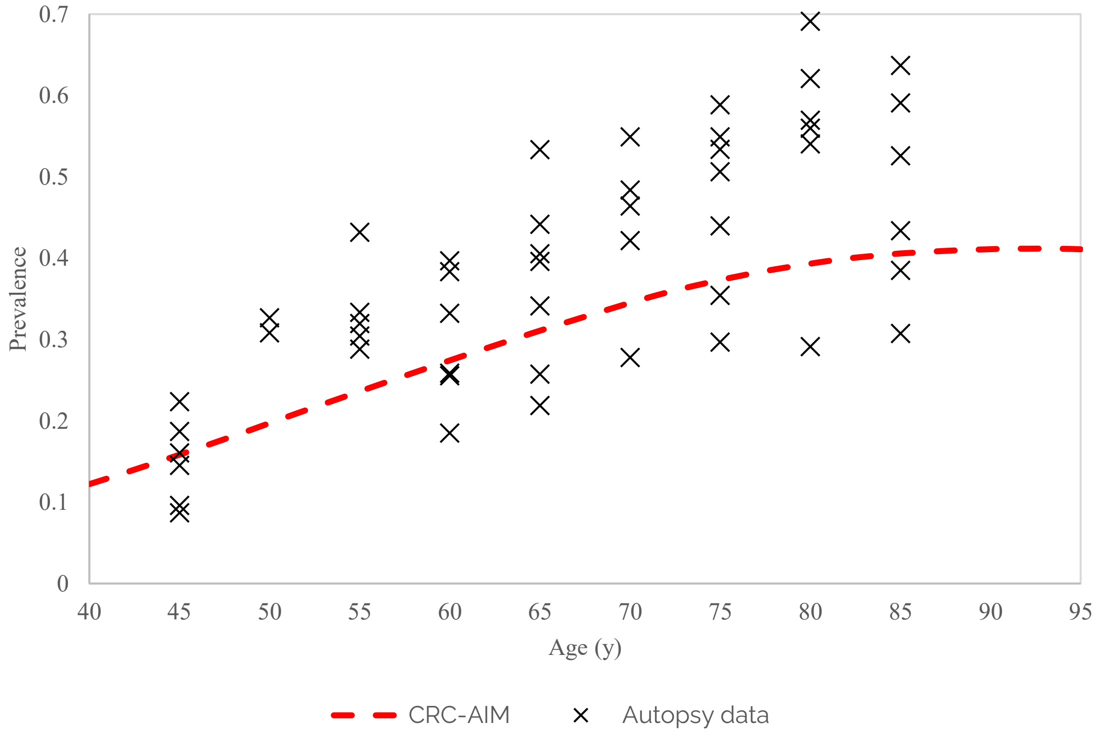


Abbreviations: CRC-AIM, Colorectal Cancer-Adenoma Incidence and Mortality model; y, year.

**Supplemental Figure 4. CRC-AIM and CISNET predictions of dwell and sojourn times(adapted from Knudsen et al.[37])**

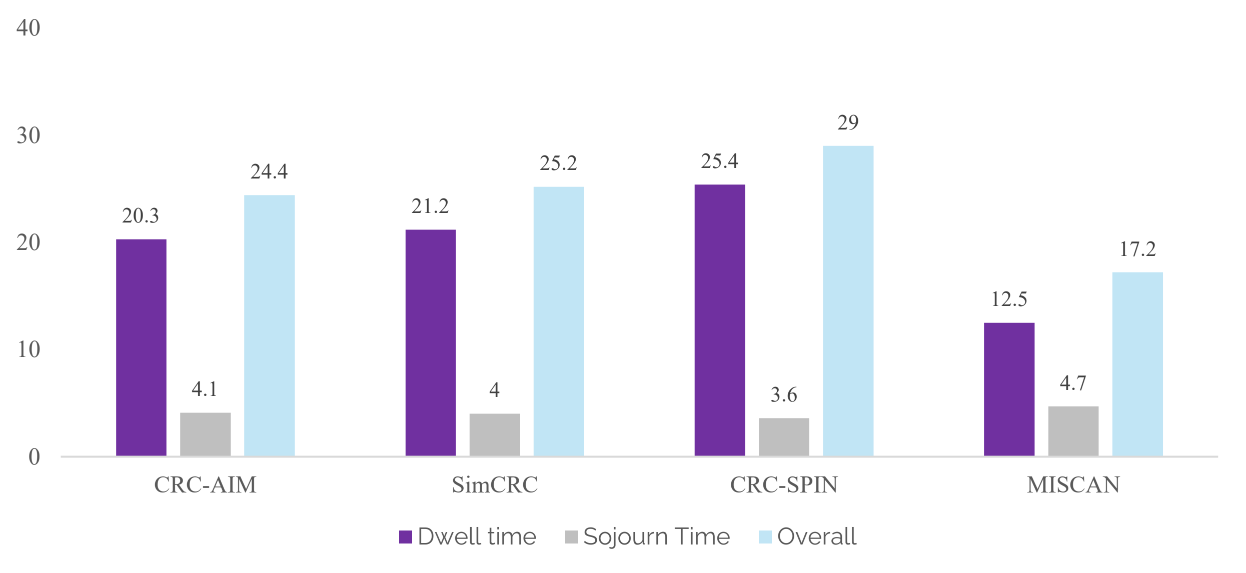


Abbreviations: CRC-AIM, Colorectal Cancer-Adenoma Incidence and Mortality model; CRC-SPIN, ColoRectal Cancer Simulated Population Incidence and Natural history model; MISCAN, MIcrosimulation SCreening Analysis; SimCRC, Simulation Model of Colorectal Cancer.

**Supplemental Table 6. Secondary calibration targets point estimates, tolerance intervals, and CRC-AIM estimates**

| Study | Target | Point estimate | Lower bound | Upper bound | CRC-AIM |
|---|---|---|---|---|---|
| Imperiale et al.[32] | Detected preclinical cancers per 1000 people | 6.0 | 1.5 | 15.9 | 4.97 |
| Lieberman et al.[33] | Preclinical CRCs per 1000 lesions, 6–9 mm | 2.5 | 0.0 | 18.0 | 5.92 |
| | Preclinical CRCs per 1000 lesions, ≥ 10 mm | 32.1 | 13.0 | 64.1 | 23.21 |
| Church[34] | Preclinical CRCs per 1000 lesions, [6, 10) mm | 2.4 | 0.0 | 29.5 | 9.35 |
| | Preclinical CRCs per 1000 lesions, ≥ 10 mm | 42.3 | 15.6 | 88.7 | 45.37 |

* Upper and lower bounds were based on the exact confidence interval. 99.99% confidence interval was used due to high level of uncertainty.

Abbreviations: CRC, colorectal cancer; CRC-AIM, Colorectal Cancer-Adenoma Incidence and Mortality model.

### E. CRC-AIM Screening Cross-model validation

Results from three CISNET models (CRC-SPIN, MISCAN-COLON, and SimCRC), were included in the 2021 USPSTF CRC screening guideline update,[37] and used to cross validate the CRC-AIM model. Model outcomes were compared including life-years gained (LYG) with screening, CRC incidence and deaths (in the presence and absence of screening), and total number of colonoscopies conducted by screening modality. Three screening strategies (at their recommended screening intervals) for individuals aged 45 to 75 years were compared: colonoscopy every 10 years, annual fecal immunochemical test (FIT), and triennial multi-target stool DNA (mt-sDNA) test. Sensitivity for stool-tests were calibrated to match the overall non-advanced adenoma sensitivity (consistent with the USPSTF modeling approach).[37] With assumed perfect screening adherence (100%), an incidence rate ratio was applied to reproduce the increasing underlying risk of CRC development after 1970.[36] CRC incidence for adults aged <50 years who are not screening eligible has substantially increased for both men and women in colon and rectum.[36] Previous analysis from USPSTF,[37] report an incidence rate ratio set to 1.19. This incidence rate ratio was assumed to be an increase in the baseline log-risk, ($\beta_0$) of adenoma and applied through each simulated individual's lifespan.

Life-years gained (Supplemental Figure 5), as well as incidence (Supplemental Figure 6) and mortality(Supplemental Figure 7) reductions associated with colonoscopy, FIT, and mt-sDNA screening strategies, were estimated by CRC-AIM and compared to CISNET model predictions.

**Supplemental Figure 5. CRC-AIM and CISNET[37] estimates of life-years gained from screening by modality (adapted from Vahdat et al.[26])**

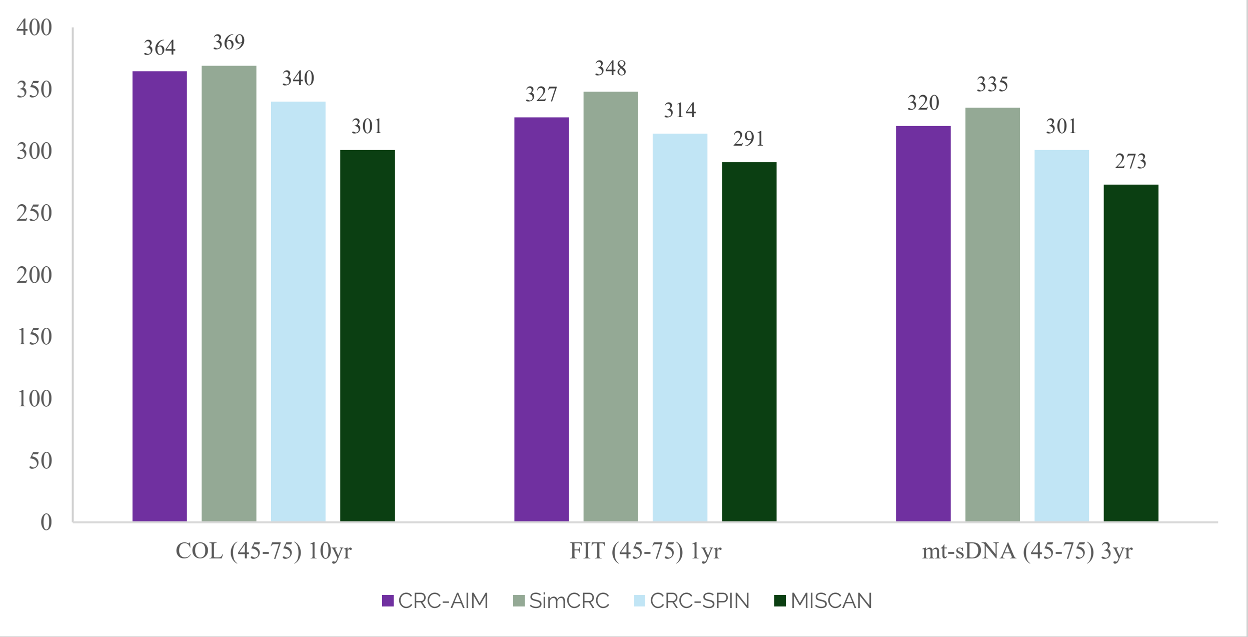


Abbreviations: COL, colonoscopy; CRC-AIM, Colorectal Cancer-Adenoma Incidence and Mortality model; CRC-SPIN, ColoRectal Cancer Simulated Population Incidence and Natural history model; FIT, fecal immunochemical test; MISCAN, MIcrosimulation SCreening Analysis; mt-sDNA, multi-target stool DNA; SimCRC, Simulation Model of Colorectal Cancer; yr, year.

**Supplemental Figure 6. CRC-AIM and CISNET estimates of colorectal cancer incidence reduction by screening modality (adapted from Knudsen et al.[37])**

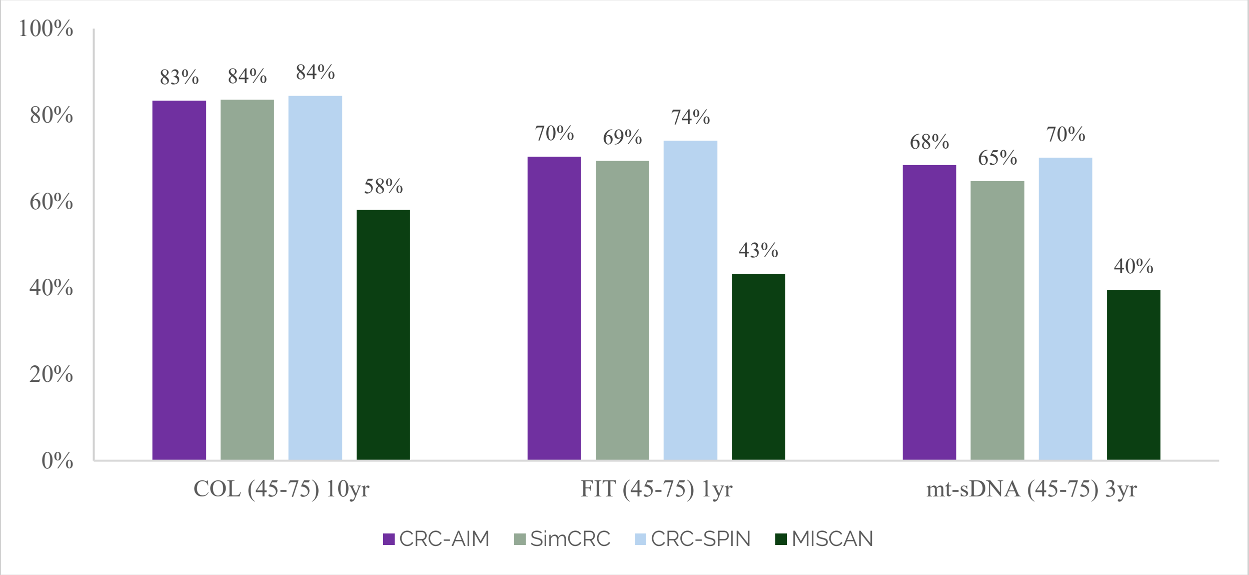


Abbreviations: COL, colonoscopy; CRC-AIM, Colorectal Cancer-Adenoma Incidence and Mortality model; CRC-SPIN, ColoRectal Cancer Simulated Population Incidence and Natural history model; FIT, fecal immunochemical test; MISCAN, MIcrosimulation SCreening Analysis; mt-sDNA, multi-target stool DNA; SimCRC, Simulation Model of Colorectal Cancer; yr, year.

**Supplemental Figure 7. CRC-AIM and CISNET estimates of colorectal cancer mortality reduction by screening modality (adapted from Knudsen et al.[37])**

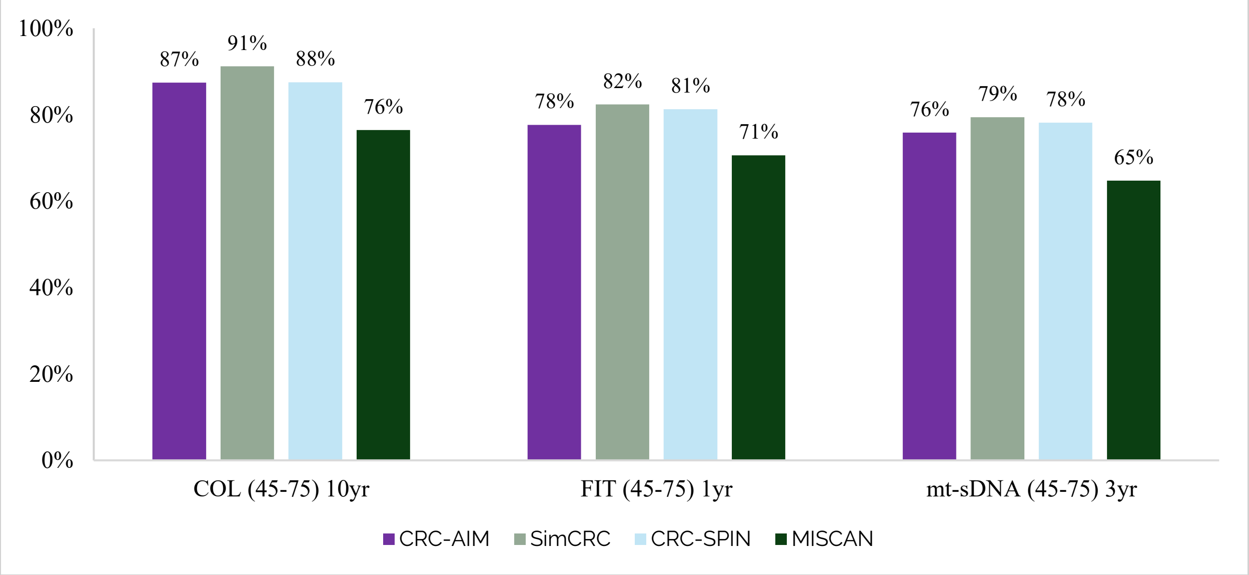


Abbreviations: COL, colonoscopy; CRC-AIM, Colorectal Cancer-Adenoma Incidence and Mortality model; CRC-SPIN, ColoRectal Cancer Simulated Population Incidence and Natural history model; FIT, fecal immunochemical test; MISCAN, MIcrosimulation SCreening Analysis; mt-sDNA, multi-target stool DNA; SimCRC, Simulation Model of Colorectal Cancer; yr, year.

### F. Overview of external validation using the United Kingdom Flexible Sigmoidoscopy Screening Trial (UKFSST)

Modeled outcomes from CRC-AIM were compared to outcomes from UKFSST (a randomized controlled trial reporting CRC incidence and mortality outcomes after one-time flexible sigmoidoscopy).[45-47] UKFSST was performed in a population that was not yet routinely screened for CRC, thus, trial results provided unique information on the preclinical duration of CRC and the impact of screening on CRC risk. As a result, many simulation models including CSNET CRC models used UKFSST as an external-validation target.[48-51]

UKFSST participants (aged 55-64 years) were randomized into control and sigmoidoscopy screening groups. Given that the UKFSST was a U.K. based trial, 1996-1998 U.K. life tables were used to adjust other-cause mortality in CRC-AIM,[52] with no other modifications to the natural history of the model. The trial was simulated 500 times, each generating a cohort with size, age, and sex distributions comparable to the observed data from the trial. Primary outcomes included hazard ratios of CRC incidence and mortality, while secondary outcomes included cumulative incidence and mortality over 17-years of follow-up. Hazard ratios of CRC incidence and CRC mortality at 17 years follow-up (Supplemental Figure 8) and cumulative probabilities of CRC incidence and mortality (Supplemental Figure 9) estimated by CRC-AIM were consistent with the reported outcomes from UKFSST,[45] establishing the external validity of CRC-AIM.

**Supplemental Figure 8. External validation with UKFSST: Hazard ratios of colorectal cancer incidence and mortality between screening and control groups over the 17 year follow-ups (adapted from Knudsen et al.[37])**

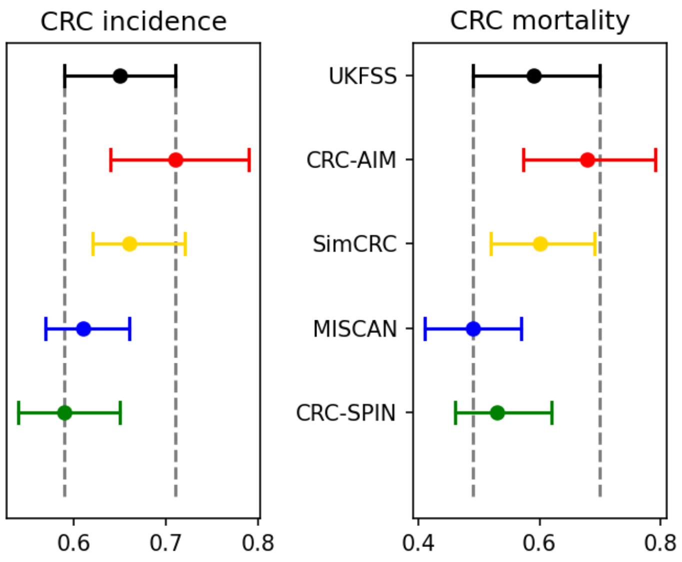


Abbreviations: CRC, colorectal cancer; CRC-AIM, Colorectal Cancer-Adenoma Incidence and Mortality model; CRC-SPIN, ColoRectal Cancer Simulated Population Incidence and Natural history model; MISCAN, MIcrosimulation SCreening Analysis; SimCRC, Simulation Model of Colorectal Cancer; UKFSST, United Kingdom Flexible Sigmoidoscopy Screening Trial.

**Supplemental Figure 9. Cumulative probabilities of CRC incidence and mortality by intervention group over 17 years of follow up (adapted from DeYoreo et al.[53])**

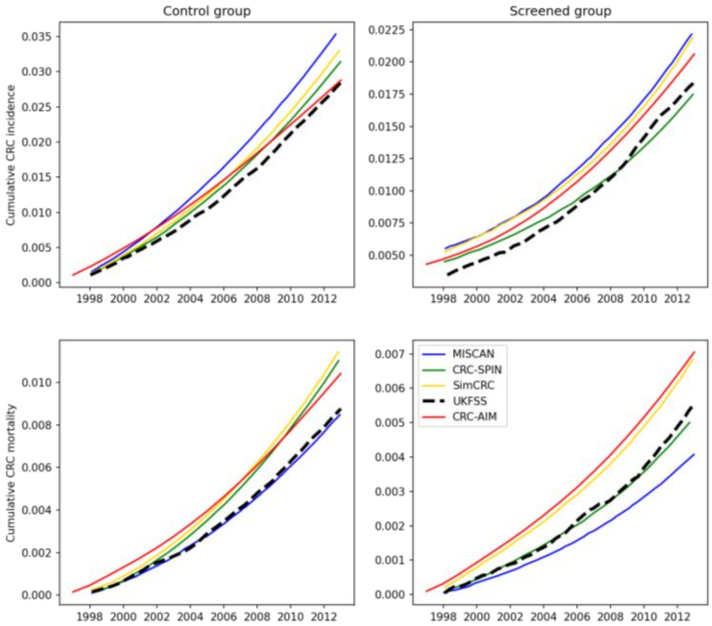


Abbreviations: CRC, colorectal cancer; CRC-AIM, Colorectal Cancer-Adenoma Incidence and Mortality model; CRC-SPIN, ColoRectal Cancer Simulated Population Incidence and Natural history model; MISCAN, MIcrosimulation SCreening Analysis; SimCRC, Simulation Model of Colorectal Cancer; UKFSST, United Kingdom Flexible Sigmoidoscopy Screening Trial.
